# Procedural time reduction associated with active esophageal cooling during pulmonary vein isolation

**DOI:** 10.1101/2021.10.04.21264525

**Authors:** Chris Joseph, Jacob Sherman, Alex Ro, Wes Fisher, Jose Nazari, Mark Metzl

## Abstract

**Background:** Active esophageal cooling is increasingly utilized as an alternative to luminal esophageal temperature (LET) monitoring for protection against thermal injury during pulmonary vein isolation (PVI) when treating atrial fibrillation (AF). Published data demonstrate the efficacy of active cooling in reducing thermal injury, but impacts on procedural efficiency are not as well characterized. LET monitoring compels pauses in ablation due to heat stacking and temperature overheating alarms that in turn delay progress of the PVI procedure, whereas active esophageal cooling allows avoidance of this phenomenon.

**Objective:** Measure the change in PVI procedure duration after implementation of active esophageal cooling as a protective measure against esophageal injury.

**Methods:** We performed a retrospective review under IRB approval of patients with AF undergoing PVI between January 2018 to February 2020. For each patient, we recorded age, gender, and total procedure time. We then compared procedure times before and after the implementation of active esophageal cooling as a replacement for LET monitoring.

**Results:** A total of 373 patients received PVI over the study period. LET monitoring using a multi-sensor probe was performed in 198 patients, and active esophageal cooling using a dedicated device was performed in 175 patients. Patient characteristics did not significantly differ between groups (mean age of 67 years, and gender 37.4% female). Mean procedure time was 146 minutes in the LET monitored patients, and 110 minutes in the actively cooled patients, representing a reduction of 36 minutes, or 24.7% (p<.001). Median procedure time was 141 minutes in the LET monitored patients and 100 minutes in the actively cooled patients, for a reduction of 41 minutes, or 29.1% (p<.001).

**Conclusions:** Implementation of active esophageal cooling for protection against esophageal injury during PVI was associated with a significantly large reduction in procedure duration.

## Introduction

As the utilization of catheter ablation for the treatment of atrial fibrillation increases, a focus on thermal injury has increased, given the risks associated with radiofrequency ablation.[1] Until the recent introduction of active cooling, luminal esophageal temperature (LET) monitoring had been the standard of care.[2] However, LET often notifies the electrophysiologist after the esophageal temperature has reached dangerous levels—after injury has occurred.[3, 4] Consequently, temperature alarms in ablations that utilize LET can result in frequent pauses to wait for luminal temperature to return to safe levels. These pauses lead to increased procedure times and suboptimal ablations given an increase in the continuity index of each ablation.[5, 6] Reducing procedure time can also reduce complications such as postoperative cognitive dysfunction.[7]

Active cooling appears to reduce the risk of severe esophageal injury.[8-10] To date, despite thousands of uses of active cooling (currently over 6000), no atrioesophageal fistula has yet been reported with active esophageal cooling.[11] Hypothesized mechanisms for this effect extend beyond dissipation of heat, and include the prevention of esophageal wall temperatures from reaching lethal isotherm temperatures and the mitigation of the inflammatory cascade contributing to burn progression.[12-15] Because active cooling eliminates overheating and avoids temperature alarms, active cooling also allows electrophysiologists to operate without pauses and unnecessary time delays.[9, 15, 16] In order to quantify this effect, we measured procedure lengths at our single center two-hospital system and compared procedure times before and after the introduction of active esophageal cooling as a means for reducing esophageal thermal injury.

## Methods

### Study Design and Patient Selection

This study was a retrospective review under IRB approval of all patients with atrial fibrillation who were treated with left atrial RF ablation by two physicians during the period January 2018 to February 2020. No patients fitting these criteria were excluded. Patients having first-time or redo ablations were included, and AF types included paroxysmal, persistent, and long-standing persistent.

### Data Collection and Definition

For each patient, we obtained and recorded the total procedure time as recorded in the electronic medical record or existing practice records maintained by the physician practice group. Total procedure time was defined as the time from the “time out” procedure marking the beginning of the case, to the time of sheath removal.

### Ablation procedure

Two electrophysiologist physicians performed primarily wide area circumferential pulmonary vein isolation with additional posterior wall isolation as needed. The posterior wall was isolated using a combination of roof and floor linear lesions, along with additional lesions to further segment the posterior wall to achieve entrance and exit block (using output of 20 mA and 5 msec pulses). Patients received general anesthesia for the ablation procedure. Anticoagulation was administered prior to ablation with a heparinized target ACT of greater than 300 seconds. In the right femoral vein, a transseptal catheter, decapolar coronary sinus catheter (Webster CS Bidirectional, Biosense Webster, Inc., Diamond Bar, CA, USA), and intracardiac echocardiographic (ICE) catheter (Soundstar, Biosense Webster, Inc., Diamond Bar, CA, USA) were placed. A very low to no flouroscopy protocol was followed for all procedures. A 3D geometry was created using the Carto system (Biosense Webster, Inc., Diamond Bar, CA, USA). A single transseptal puncture was performed under ICE and electroanatomic mapping guidance. Electroanatomic mapping, vein voltage, and pace mapping was performed using a multipolar mapping catheter (PentaRay, Biosense Webster, Inc., Diamond Bar, CA, USA).

For ablation, an externally irrigated ablation catheter (ST/SF™, Biosense Webster, Inc., Diamond Bar, CA, USA) was used in all cases. The pulmonary veins were isolated by delivery of RF applications circumferentially to the antral regions to produce a minimum of entrance and exit block for at least 20 minutes. A Smartablate™ generator (Biosense Webster, Inc.™, Diamond Bar, CA, USA) was used to deliver RF energy, with a setpoint of 50 W on all patients and all areas of the left atrium. The Visitag Surpoint™ module (Ablation index) was utilized during ablations, with a target of 400 units on the posterior wall, and 550 units on the anterior wall, lateral wall, and septum and an intertag distance of less than 6 mm. Catheter tip temperature, power, and impedance were recorded for each RF energy application.

### Esophageal protection

Prior to implementing the use of active cooling, LET monitoring was utilized using a multi-sensor probe (Circa S-Cath™, Circa Scientific, Inc., Englewood, CO, USA). RF ablation was stopped and the site of ablation moved to a different distant area of the veins for any alarm over 0.2°C/sec, or a temperature exceeding 38.5°C. After the adoption of active cooling, ablation proceeded in a point-to-point fashion uninterrupted by pauses or alarms. Except for the change to the esophageal cooling protocol in the treatment group, the ablation procedure for patients in both groups was the same.

### Statistical analysis

Data were analyzed with IBM SPSS Statistics version 26 (IBM, Armonk, NY), with descriptive statistics (mean, median, and standard deviations), and comparison between groups with the Mann-Whitney U test reported.

## Results

### Baseline characteristics

A total of 373 patients received PVI over the study period. LET monitoring using the multi-sensor probe was performed in 198 patients, and active esophageal cooling was performed in 175 patients. Patient characteristics did not significantly differ between groups. The mean age of LET-monitored patients was 67 years (SD 11 years), while the mean age of actively cooled patients was 69 years (SD 10 years). Patient gender was 36.9% female in the LET-monitored group, and 39.2% female in the actively cooled group.

### Association between active esophageal cooling and procedure time

Mean procedure time was 146 minutes in the LET monitored patients, and 110 minutes in the actively cooled patients, representing a reduction of 36 minutes, or 24.7% (p<.001). Median procedure time was 141 minutes in the LET monitored patients and 100 minutes in the actively cooled patients, for a reduction of 41 minutes, or 29.1% (p<.001). Figure 1 shows a histogram of procedure times for each of the two cohorts (LET monitored and actively cooled).

**Figure 1.**
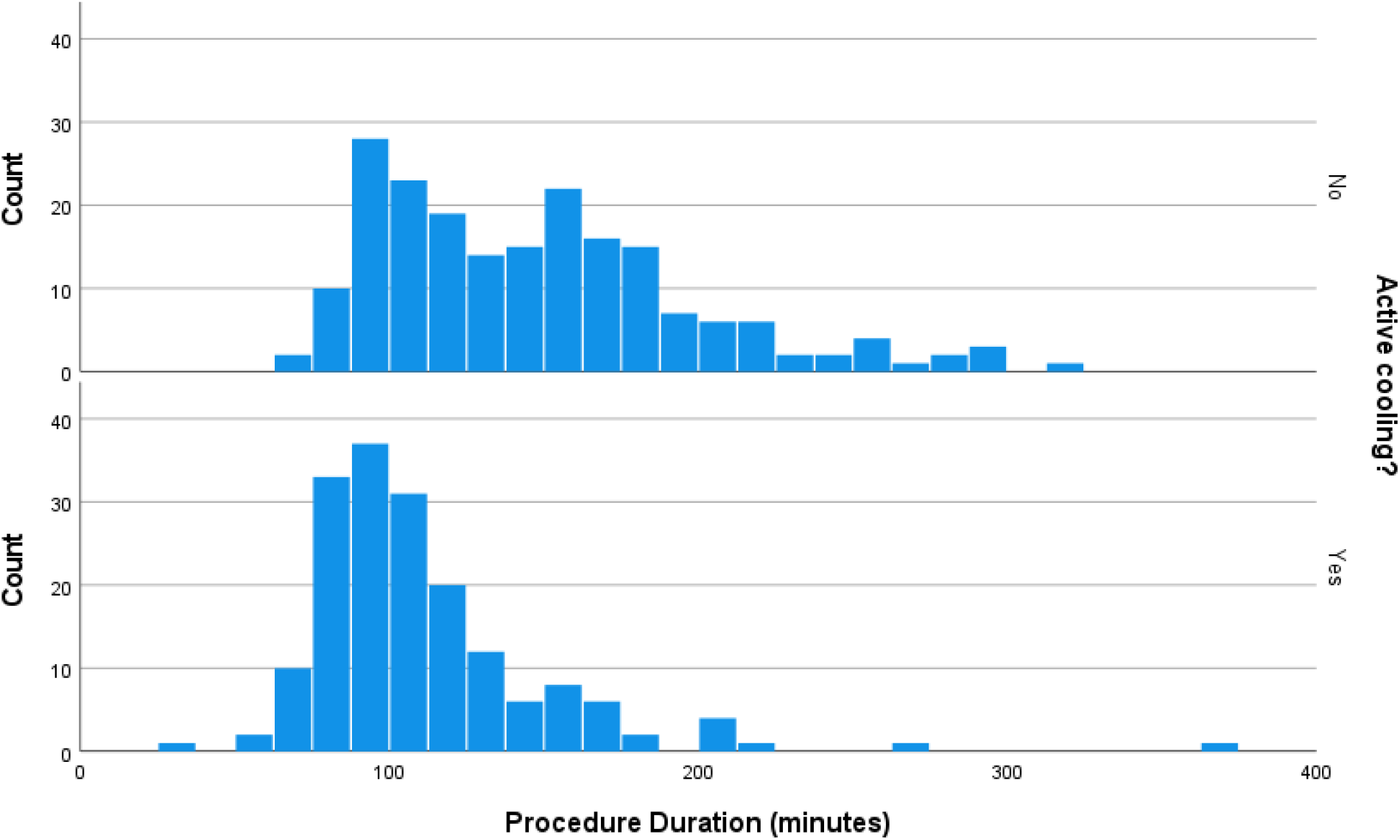
Histogram of procedure times for each of two cohorts totaling 373 patients (198 received LET monitoring using a multi-sensor probe, and 175 received active esophageal cooling). The top histogram depicts procedure times for LET-monitored patients, and bottom histogram depicts procedure times for actively cooled patients.

## Discussion

In this largest study to date on the impact of active esophageal cooling on procedure duration, a significant reduction in time was evident after the implementation of active cooling as an esophageal protective strategy. A mean procedure time reduction of 36 minutes was seen, representing a 24.7% reduction from baseline, and a median procedure time reduction of 41 minutes was seen, representing a 29.1% reduction from baseline. In both measures of procedure duration, a procedure time of under 2 hours was attained after converting to active esophagealcooling from LET monitoring during PVI cases (mean of 110 minutes, and median of 100 minutes).

Procedure times for PVI with RF ablation will naturally vary by operator and site, but recent studies have reported times of 132 to 188 minutes for RF ablations utilizing a variety of esophageal protection techniques (single-sensor LET monitoring, multi-sensor LET monitoring, or power reduction on the posterior wall).[17-20] As such, the reductions seen in this study are notable. The likely mechanism for this reduction stems from the elimination of pauses in ablation that are compelled by LET sensor alarms and concerns over heat stacking. Alarm sensitivity can be adjusted on some systems, raising or lowering the sensitivity of measurements. The settings utilized during LET monitoring prior to the adoption of active esophageal cooling were such that alarms were triggered, and RF ablation was stopped (with the site of ablation moved to a different distant area) at a temperature rise of 0.2°C/second or greater, or a temperature exceeding 38.5°C. Although these are fairly typical settings, higher thresholds have been used by other groups, which in turn may influence results (reducing the number of pauses required, at the cost of increased risk of esophageal injury).[18]

Despite shortcomings in protective effects, LET continues to be the standard of care in many electrophysiology labs across the world. Recent studies, including randomized controlled and prospective interventional studies have raised concerns that LET monitoring does not reduce, and may actually increase, esophageal lesion formation.[17, 18, 21] Proposed mechanisms for this finding involve physical limitations in adequately positioning temperature sensors to detect thermal insults, and inherent limitations in detecting temperature rise before damage has occurred.[3, 4] In contrast, three randomized, controlled studies have demonstrated benefits with active esophageal cooling, with the largest study of 120 patients showing reductions of all lesion formation of 83%, and reductions of severe lesion formation of 100% on per-protocol analysis.[2, 8, 22, 23] Shortening of the procedure time may provide further incentive to transition from a passive monitoring strategy to an active cooling strategy for esophageal protection. In addition to reducing lab costs and improving staff satisfaction, shorter procedure times may also reduce postoperative complications such as postoperative cognitive dysfunction.[7] Beyond the acute benefits, long-term efficacy improvement has also been suggested to occur with active cooling, with trends towards greater freedom from AF at 12 months after active cooling than with LET monitoring.[24] This is hypothesized to be due to the lower Continuity Index that can be obtained with active cooling, allowing consistent, contiguous placement of lesions without the pauses and repositioning required with LET monitoring.[6]

A recent study found a 35% reduction in fluoroscopy usage after the implementation of active cooling (compared to patients treated with single-sensor LET monitoring).[16] Zagrodzky et al. did not report shorter procedure times, but their procedure durations were sufficiently short that further significant shortening may have been unachievable.

## Limitations

While this study demonstrates an association between active cooling and reduced overall procedure time, we are unable to definitively conclude that the reduction of procedure time is due to the use of active cooling. No significant procedural changes were made during the period of observation other than the switch to active esophageal cooling from LET monitoring, which lends support to a causative effect from active esophageal cooling. We were not able to quantify the number of pauses that occurred during ablations utilizing LET monitoring, since these are typically not captured in the medical record. Additionally, given the use of multi-sensor temperature probes in this study, the aforementioned procedure time benefit may not be applicable to labs that utilize single sensor probes, although this is not likely to impact procedure time significantly.

## Conclusion

Implementation of active esophageal cooling for protection against esophageal injury during PVI was associated with a significant and large reduction in procedure duration

## Data Availability

Data are available upon reasonable request.

## Acknowledgements

Marisa Durante, Division of Cardiology Research Study Coordinator, NorthShore and Erik Kulstad, MD, UT Southwestern Medical Center and Attune Medical.

